# Identifying patients presenting in pain to the adult emergency department: A binary classification task and description of prevalence

**DOI:** 10.1101/2022.05.29.22275652

**Authors:** J.A. Hughes, C. Douglas, L. Jones, N.J. Brown, A. Nguyen, R. Jarugula, A. Lyrstedt, S. Hazelwood, Y. Wu, F. Saleh, K. Chu

**Author notes:** Contribution to the field. Pain is the most common symptom that patients will experience on presentation to the emergency department (ED). Current figures suggest that anywhere from 52% to 85% of all patients presenting to the ED will have pain on arrival. This equates to between 4.57 and 7.48 million patients presenting with pain to the ED each year in Australia alone. Current literature reports that EDs are poor at treating pain with an over-reliance on the use of opioids. Current methods for measuring and estimating this population are poor with much of the information coming from small samples in single-site studies. This work presents a rigorous method for retrospectively identifying patients presenting in pain from the ED record over three years, describing the previlance, demographics and treatment. This type of epidemiological work is required if pain care is to be improved in the ED. This work represents the first stage of a program of research to utilise artificial intelligence to identify patients presenting in pain to the ED at a population-based level, information that is required if we are to make sustainable, systems-level changes to how pain is treated in the ED.

## Abstract

**Background:** Accurate, reliable and efficient measures of pain-related presentations are essential to evaluate and improve pain care in the ED. Estimates of pain prevalence on arrival to the emergency department (ED) vary depending on the methods used. Artificial intelligence (AI) approaches are likely to be the future for identifying patients in pain from electronic health records (EHR). However, we need a robust method to identify these patients before this can occur. This study aims to identify patients presenting in pain to the ED using binary classification and to describe the population, treatment and outcomes.

**Methods:** This study employs a cross-sectional design using retrospective data routinely collected in the EHR at a single ED. A random sample of 10 000 patients was selected for inclusion over three years. Triage nursing assessment underwent binary classification by three expert clinicians. The prevalence of pain on arrival is the primary outcome. Patients with pain were compared to those without pain on arrival regarding demographics, treatment and outcomes.

**Results:** The prevalence of pain on arrival was 55.2% (95%CI 54.2% - 56.2%). Patients who presented in pain differed from those without pain in terms of age, country of birth, socioeconomic status, mode of arrival, urgency and discharge destination. The median time to first analgesic medication was 65min (IQR 38 – 114 min), and 45.6% (95% CI 44.3% - 46.9%) of patients arriving in pain received analgesic medication.

**Conclusions:** The prevalence of pain on arrival compares well with previously reported figures using similar methods. Differences in the cohort presenting in pain compared to the population may represent differences in the prevalence or be an extension of previous bias seen in the documentation of pain. This work has set a rigorous methodology for identifying patients presenting with pain from the EHR. It will form the basis for future applications of AI to identify patients presenting in pain to the ED.

## Introduction

Pain has been reported as the most common presenting symptom to the emergency department (ED) worldwide (1, 2). The considerable between-study variation in prevalence suggests that it is difficult to accurately identify patients with pain in ED populations. Commonly reported figures range from 49% to 78% (3, 4). Some variation is explained by methodology, with retrospective data yielding a lower prevalence (1, 3, 5-9) than directly asking patients upon presentation (4, 10-13). However, pain prevalence studies lack consistency, and many methods require significant resource allocation for small sample sizes that may not generalise to target populations.

Interventions aimed at improving pain care in the ED have relied on clinicians to document self-reported pain intensity on presentation (2, 14) or selected painful diagnoses at discharge (15) to identify the population. These methods may underestimate the prevalence of pain on arrival to the ED and introduce misclassification bias. Without a straightforward, reproducible method for identifying the prevalence of pain in the ED, a description of the characteristics of this population, treatment and outcomes, significant advances in improving pain care may be limited. In this work, we describe an adaptation of a retrospective method to identify patients presenting in pain to the ED. We then describe the population patient characteristics and pharmacological treatment provided. This work represents a first step to developing an Artificial Intelligence (AI) method of identifying pain retrospectively form electronic health records (EHR).

## Background

It is widely accepted that pain is the most common symptom patients experience when presenting to the ED for care (2, 5, 12, 15-17). While it is not uncommon to see figures between 49% to 78% (3, 4), more commonly, authors cite figures in the range of 60% - 70% of all patients who present to the ED (1, 16, 17). The majority of publications on ED pain care rely on just one reference to support this statement (1). This finding is based on one week’s data from an urban ED in the USA collected almost 20 years ago.

Table 1 outlines 16 pain prevalence studies in the ED from eight countries using three distinct approaches over the last 24 years. Retrospective chart review (RCR) was used in most studies to identify specific words, phrases, assessments, diagnoses, or treatments documented in routine care that indicated that the patient had presented in pain (1, 3, 5-9, 18). Prospective self-report (PSR) of pain on arrival was used in five of the identified studies (4, 10-13), usually conducted by a dedicated researcher located in the ED during defined periods. The third method involved completing voluntary post-hoc patient experience (Exp) surveys administered by large health systems (19-21). These surveys were completed up to three months post-discharge from the ED and relied on patients to return the surveys. The prevalence reported were grouped by the methods used for the identification of pain. The RCR group (49.3% - 62.3%) (3, 7) and the Exp group were similar in their estimates of prevalence (51.0% - 67.0%) (19, 21). However, the PSR group consistently estimated the prevalence of pain on arrival 11.8% - 21.4% higher than the RCR and Exp groups at 70.7% - 78.8% (4, 10).

**Table 1:** Studies identifying the prevalence of pain experienced in the adult Emergency Department

| Study | Method | Setting | Identification of patient in pain | Prevalence |
| --- | --- | --- | --- | --- |
| **Retrospective Chart Review (RCR)** |  |  |  |  |
| Cordell et al. (2002)<br>United States of America | Retrospective chart review of all patients presenting over seven consecutive days. | Urban, tertiary care referral centre with an annual census of 90 000 presentations | Pain or pain equivalent word used in the clinical notes associated with presentation (list provided) | 61.2% (1019/1665)<br>(95%CI 58.9% - 63.5%) |
| Grant (2006)<br>United Kingdom | Secondary analysis of two non-consecutive months worth of presentations over the age of 18 years. | “Busy” ED that was part of the NHS. | Any patient who had any form of pain as part of their presenting complaint, symptom or diagnosis: or patients who received any form of analgesia | 25.9% (473/1821)<br>(95%CI 24.0% - 28.0%) |
| Wong et al. (2011)<br>Hong Kong | Retrospective review of a systematic sample of seven consecutive days of presentations over the age of 5 years | A regional trauma centre in Hong Kong with an annual attendance of 156 000 presentations | Any patient who had “pain” or “pain equivalent words” documented on their attendance record had analgesia administered. | 60.7% (300/494)<br>(95%CI 56.3% - 64.9%) |
| Gueant et al. (2011)<br>France | Patients over the age of 15 years were prospectively selected. Pain scores were recorded for patients selected as part of the generation of the medical record. | 50 EDs (both public and private) across France | Documentation of a pain score greater than 0/10. | 62.3% (7265/11670)<br>(95%CI 61.4% - 63.1%) |
| Chang et al. (2014)<br>United States | Retrospective review of 10 years of national cross-sectional data on ED visits of patients 18 years and older without malignancy | Nationwide – United States of America | Patient-reported symptoms that indicated pain or a physician diagnosis of pain based on the ICD-9 code. | 52.2% - 57.3%* |
| Jones et al. (2017)<br>New Zealand | Retrospective chart review of randomly selected presentations | 26 New Zealand EDs over 6 years | Any painful condition | 56.0% (866/1547)<br>(95%CI 53.5% - 58.4%) |
| Thornton et al. (2018)<br>United Kingdom | Retrospective review of seven days of consecutive presentations | A single large NHS ED | Pain score documented greater than 0 | 49.3% (573/1163)<br>(95%CI 46.5% - 52.2%) |
| Hughes et al. (2021)<br>Australia | Retrospective review of triage nursing assessment by one trained abstractor | A single large inner-city ED with an annual attendance of 80 000. | Pain or pain equivalent word used in the Triage nursing assessment on presentation. | 50.8% (1015/2000)<br>(95% 48.6% - 52.9%) |
| **Prospective Self Report (PSR)** |  |  |  |  |
| Johnson et al. (1997)<br>United States of America | Prospective recruitment of every attendance at two hospitals by a research assistant over 12 hours a day for seven consecutive days | Two university-affiliated teaching hospital EDs, one general and one paediatric. | Self-report of pain on arrival | 71.2% (200/281)<br>(95%CI 65.4% - 76.3%)+ |
| Tanabe et al. (1999)<br>United States of America | Prospective recruitment of patients over the age of 18 years for an interview regarding their pain. | Large urban ED with annual attendances of 35 000 per year | Self-report of pain using verbal descriptive scale and numerical pain rating scale | 78.8% (160/203)<br>(95%CI 72.7% - 83.9%) |
| Karwowski-Soulie et al., (2006)<br>France | A structured questionnaire administered by a research assistant 24hours a day over 15 consecutive days | ED of a university hospital seeing approximately 35 000 presentations per year. | Self-report of pain using either a numerical descriptor scale or verbal pain intensity scale. | 77.5% (563/726)<br>(95%CI 74.4% - 80.4%) |
| Mura et al. (2017)<br>Italy | Prospective observational study where every patient was asked at the moment of triage. | Second-level urban ED | Self-report of a numerical pain rating scale | 70.7% (732/1035)<br>(95%CI 67.9% - 73.4%) |
| Minguez Maso et al., (2013)<br>Spain | Cross-sectional study where a physician asked patients older than 15 years about the presence of pain over seven consecutive days | University hospital with 87 000 presentations per year | Self-report of pain | 75% (501/668)<br>(95%CI 71.6% - 78.1%) |
| **Patient Experience Surveys (Exp)** |  |  |  |  |
| Queensland Health (2015)<br>Australia | Patient experience survey completed over three months utilising computer-assisted telephone interviewing post-discharge from the ED. | 53 EDs of all sizes and locations in Queensland | Post-hoc self-report of pain. Question - Were you ever in any pain while in the ED? | 59.0% (8618/14607)<br>(95%CI 58.2% - 59.8%) |
| Care Quality Commission (2018) United Kingdom | Patient experience survey completed over one month. Survey delivered by post after discharge from an ED. | The EDs of 132 NHS trusts | Post-hoc self-report of pain when they visited the ED | 67% (28005/41799)<br>(95%CI 66.5% - 67.4%) |
| New South Wales Health (2020) Australia | Patient experience survey. A questionnaire sent to patients who have presented into the ED approximately three months after presentation | 77 large EDs in New South Wales | Post-hoc self-report of pain. Question - Were you ever in pain while in the ED? | 51% (8839/17331)<br>(95%CI 50.3% - 51.7%) |

\* Figures were reported yearly over ten years, with no yearly denominator to calculate confidence intervals. + Both adult and paediatric figures were presented; however, only adult figures are presented here. ED – Emergency Department, CI – Confidence Interval, NHS – National Health Service, the government-supported universal health system of the United Kingdom, ICD – International Classification of Disease.

Each of the three methods for calculating prevalence introduces its own inherent bias. RCR relies on the quality of the documentation used for the review and is highly dependent on the quality of abstractors (22-26). However, RCR does not require data collection outside of that collected for routine care. The retrospective nature allows for representative sampling, as has occurred in some of the studies presented (7, 8), leading to a further reduction of bias when figures are used to estimate population prevalence. PSR of pain is considered the gold standard in identifying pain (27), but this is only true for those patients who can self-report. Several vulnerable populations cannot self-report, which may be underrepresented when used (28). Employing dedicated staff to ask patients on arrival if they have pain is expensive and time-consuming, limiting the sample that can be collected and is not feasible outside of the research or quality improvement environment (11-13). Patient experience surveys have several well-documented biases inherent in their design. The retrospective, whole of system, patient experience surveys such as those used in Australia and the United Kingdom (19-21) are open to hindsight bias (29) and recall bias (30). The questions regarding pain on arrival or during the care received are likely to be influenced by these biases and not paint an accurate picture of symptoms on arrival. While samples may be selected randomly or comprise all presentations within a designated time, the inherent self-selection in responding is likely to lead to an element of response bias (31). Despite their limitation, this form of information gathering allows the whole system to report on prevalence. While exact numbers may be open to several biases, trends over time are likely to accurately represent changes in the population.

Detailed knowledge of the epidemiology of pain presenting to the ED forms the basis of the advancement of symptom science in the ED. Designing an intervention and measuring outcomes relies on having an easily reproducible methodology for identifying the target population. To be generalisable to the large population that presents to EDs worldwide, a retrospective methodology that uses widely collected patient information is required. Previous authors have identified that AI approaches, especially Natural Language Processing (NLP) are useful for the classification of symptoms in electronic health records (32, 33). Early pilot studies of the application of this technology in identifying pain within triage nursing assessments have proved feasible and successful (34, 35). To develop such tools it is essential to first develop a “gold standard” dataset of well-defined ED patients with pain, with which to train and test the algorithms. Therefore, this article will present a retrospective methodology for identifying patients presenting to a large inner-city ED with pain using the triage nursing assessment documented during the triage process. We will then describe this population in terms of their demographics, treatment provided and outcomes over three years.

## Materials and Methods

### Aims / Objectives

The aim of this study is to identify patients presenting in pain to the ED and describe the population, their treatment and outcomes. Specific objectives include:

1. To manually classify whether people presented to the ED in pain, using consensus review of the free-form text in the triage nursing assessment by an expert panel of clinicians.
2. Compare demographics (age, sex, socioeconomic status), presentation characteristics (time of arrival, mode of arrival, urgency and diagnosis), and health service outcomes (ED length-of-stay, departure destination and 72-hour representation) between patients who did and did not present to the Emergency and Trauma Centre at the Royal Brisbane and Women’s Hospital with pain during the study period.
3. Describe the analgesic treatment of ED patients presenting in pain

### Study Design

Cross-sectional design using retrospective data routinely collected during patient care. This study was reported in line with studies conducted using observational routinely-collected health data (RECORD) statement (36).

### Setting

The Emergency and Trauma Centre (ETC) at the Royal Brisbane and Women’s Hospital (RBWH) is an adult ED at a principal referral hospital in Queensland, Australia. The ETC sees approximately 85,000 primary presentations a year and is a major trauma referral centre and the state-wide referral centre for burns. The ETC uses an innovative model of care which combines team-based assessment and clinical streaming (37). In January 2020, the hospital became a major treatment centre for the COVID-19 pandemic, which it remains until this day.

### Ethical Approval

This study received ethical approval from the Human Research Ethics Committee of the RBWH (LNR/2021/QRBW/72976) and the University Human Research Ethics Committee of Queensland University of Technology (109147). In addition, access to confidential patient-level information was provided under the *Public Health Act* (approval number: 72976).

### Study Participants

All adult patients (18 years and older at presentation) who presented to the ETC of RBWH between 1^st^ March 2018 and 28^th^ February 2021 were eligible for inclusion within the study. Patients who were identified as an inter-hospital transfer, whose triage nursing assessment was blank or were missing a unique identifier that allowed data joining, were excluded. Three years were chosen to allow for seasonal variability.

### Measures (Variables)

Variables collected for this study were guided by Symptom Management Theory (SMT) (38, 39), particularly the application of SMT to the study of pain care in the ED (16) and previous retrospective reviews of pain care in the ED (18, 40). Patient demographics, including socioeconomic status, environmental (workload), components of symptom management strategies (treatment, including pharmacological treatment) and outcomes (length of stay and representation), were collected from pre-existing data within the information systems of the ED. In addition to these variables, the full text of the triage nursing assessment was collected to identify patients in pain. A full description of each of the variables reported on is available in the supplemental material (Supplementary material 1).

#### Analgesic Medication

The definition of an analgesic medication is complex when the broad range of medications that may be prescribed to relieve pain are considered. In emergency care, a small group of analgesic medications make up most of all medications used to relieve pain. In recent work in an Australian ED, Paracetamol (Acetominophen), Oxycodone, Ibruprofen, Morphine, and Fentanyl made up 93.8% of all first-line analgesic medications used (16). However, many medications with analgesia not considered their primary indication may induce analgesia in specific conditions (41). An example of this would be Chlorpromazine, a typical antipsychotic, in headaches and migraines (42).

In the period that this study was conducted, there were 354 unique medications dispensed from the electronic dispensing system located within the ED. Therefore, identifying which medications were used for their analgesic properties is essential for the outcomes of this and future studies. We categorised the medication dispensed in the ED into four categories by expert consensus of the clinical members of the research team (JH, KC, RJ, CD and AL):

1. *Analgesic Medication:* A primary medication to relieve pain and is expected to relieve pain in a wide variety of conditions presenting to the ED. All, or almost all use of this medication in the ED is expected to be used for analgesia.
2. *Local Anaesthetic:* A medication that modulates the pain sensation and may be used topically or b) parentally either by local infiltration or systemic administration.
3. *Drugs used to treat pain for specific conditions:* Medications generally not considered analgesic medications; however, they may be an analgesic medication when used to treat a specific condition. These may be considered a) first-line agents or b) second-line medications.
4. *Non-analgesic medications*: Medications that are not used to induce analgesia

A complete list of all medications considered an analgesic for the purposes of this study is contained within Supplementary material 2.

### Data Collection

Data was collected from two sources. First, the Emergency Department Information System (EDIS) contains information on the patient demographics, presentation, time to treatment and deposition. Second, the automated medication dispensing system (Pyxis™) provides information regarding the medications dispensed to the patient while receiving ED care. Data between these two sources were linked via the unique identifier assigned to each patient receiving care in the hospital. Linkage was completed if the date and time of dispensing (Pyxis™) were between the arrival and departure time for this episode of care (EDIS). Data was not linked for non-analgesic medications.

### Missing Data

Due to the retrospective nature of the information collected, there is missing data within some variables. Missing data is identified and reported on. Only missing data in the unique identifier or triage nursing assessment caused removal of the case from the dataset. Variables with greater than 5% missing data are assessed for the randomness of this missing data via logistic regression.

### Identification of Patients Presenting in Pain (Binary Classification Task)

A random sample of 10,000 patients presenting during the study period was selected for manual classification of whether they had presented with signs or symptoms of pain or not. This was based upon their triage nursing assessment. Two experienced emergency nurses (JH and AL) independently assessed each triage nursing assessment to determine whether it contained signs or symptoms that indicated the patient presented in pain. Where the two assessments did not agree, a third experienced emergency nurse (SH) adjudicated the outcome. Those reviewers completing this task only had the triage nursing assessment presented to them to make their assessment; no other patient information was provided. Signs or symptoms of pain may include a documented pain scale (x/10) or verbal descriptor of pain (mild, moderate or severe). In addition, other words or phrases such as “burning”, “throbbing”, or “headache” may be used to indicate pain. This is consistent with previous methods used to identify patients presenting in pain (1, 9, 18). The clinicians were also encouraged to use their clinical judgement to identify pain. Before completing this task, each reviewer underwent training from the principal investigator and received standardised instructions (Supplementary material 3). The binary classification task results are presented as the prevalence of pain on arrival, interrater reliability (via the kappa statistic), and proportion of disagreement between reviewers. The reviewers were assessed for sensitivity and specificity of pain ratings using the adjudicated ratings as the gold standard, with 95% confidence intervals reported. Reviewer sensitivity and specificity was then compared using a McNemar test (43).

### Statistical Analysis

All variables collected were described as means and standard deviations for continuous data and frequencies and percentages for categorical data. The bivariate analysis was completed using t-tests for continuous variables and chi-square for categorical variables. The t-test assumptions of normality within groups and equal variance between groups was assessed using descriptive statistics and plots (44). Variation in demographics, treatment and outcomes for patients with and without pain were presented as odds ratios and their associated 95% confidence intervals. Differences in the provision of analgesic medication were also be presented as Odds ratios and their associated 95% confidence intervals. Finally, differences in the time to the first analgesic medication are presented as Hazard ratios and assessed via the log-rank test, with the proportional hazards assumption assessed graphically using log minus log plots (45). Sensitivity and Specificity analysis between reviewers will be conducted using the DTComPair package for R (46).

## Results

### Patient selection

There were 250,772 presentations to the ETC at RBWH between 1^st^ March 2018 and 28^th^ February 2021. Once exclusion criteria were applied (patients less than 18 years and inter-hospital transfers), 236,353 were eligible for inclusion. A random sample of 10,000 patients was identified using the RANDBETWEEN random number generator in Microsoft Excel. Twenty-four of these patients were missing unique identifiers that allowed joining between datasets. Therefore the final sample for this study was 9,976 presentations (see Figure 1).

**Figure 1:**
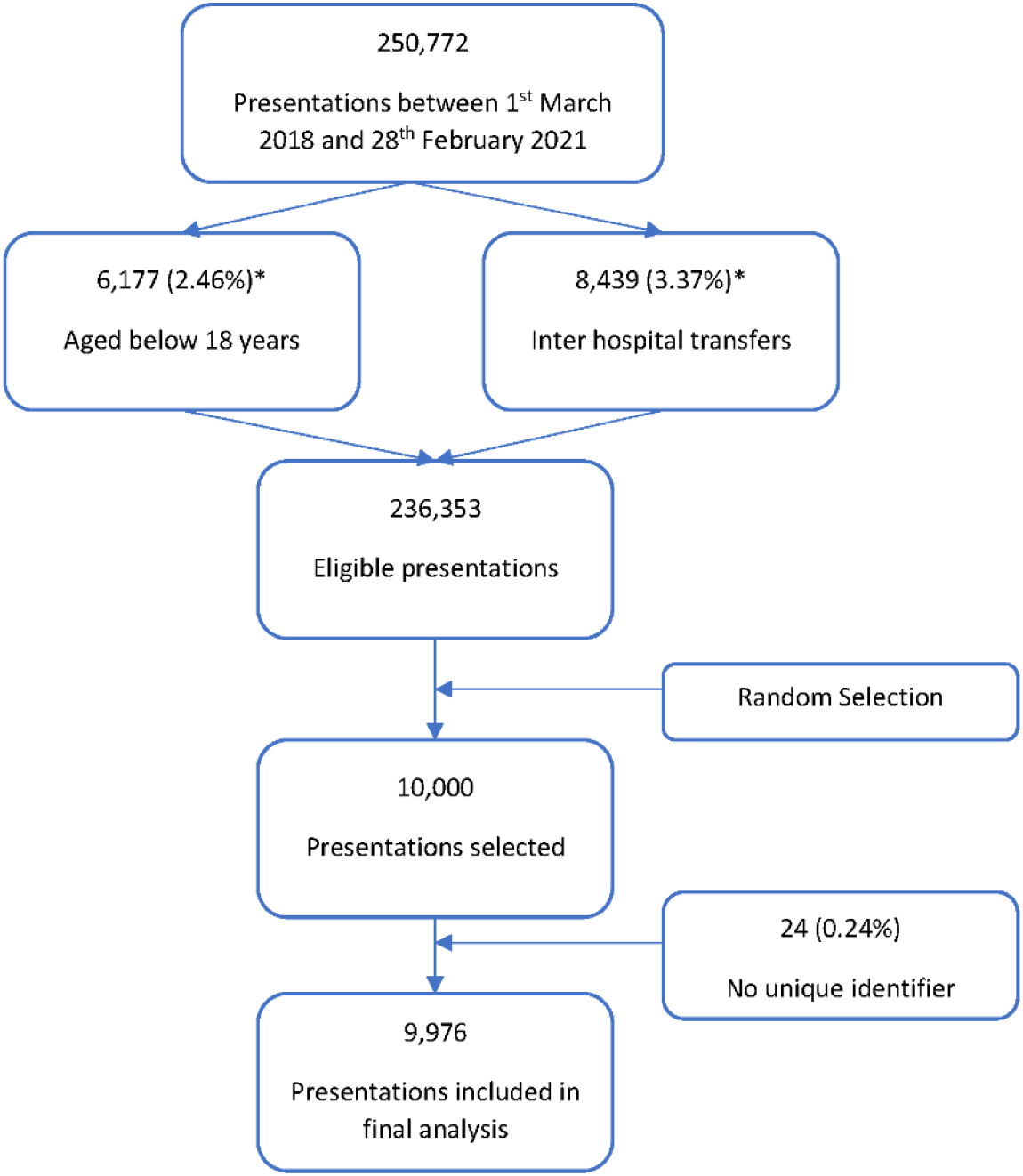
Patient selection flowchart * Presentations may have been excluded for more than one reason.

The final sample of 9,976 presentations did not differ to the population in terms of age, sex, socioeconomic status (measured by the Index of Relative Advantage or Disadvantage), mode of arrival or urgency (measured by the Australasian Triage Score). The environment where they received care also did not differ in terms of the average daily time to be seen by a medical officer, daily presentations or daily ED length of stay. These figures are presented in Appendix A.

### Missing Data

As expected for retrospective chart review, there were missing data. Five variables collected directly from the source contained missing data (sex, date of birth, postcode, Indigenous status, and time to be seen by a Provider). Except for time to be seen by a provider, all of the other variables were missing in less than 0.2% of the population. There were 600 (6.01%) missing times to be seen where there was no treating Provider. A large proportion of these can be accounted for by patients that did not wait to be seen, left against medical advice or were removed because of unacceptable behaviour (n=543, 5.44%). Patients who had this departure status were much more likely (OR 5.54, 95% CI 4.45, 6.89, p<0.001) to be missing a time to be seen.

Two variables had documented non-responses. Country of birth was “Not Stated” in 102 (1.02%) of cases, and Employment status was documented as “Not Stated” or “Unknown” in 2692 (26.98%) of cases. While these responses form unique groups, they represent information missing in the dataset.

### Identification of patients in pain

After training and written instructions, two independent reviewers categorised the sample of 9,976 presentations into either “Pain” or “No Pain”. Reviewer one identified pain in 5,475 (54.9%) cases and reviewer two identified pain in 5,312 (53.2%) cases. Reviewers one and two disagreed on the classification in 957 (9.59%) presentations referred to reviewer three (κ = 0.807 (95% CI 0.795, 0.818), z = 80.703, p <0.001). This process was undertaken over approximately twelve weeks. Both Reviewers had high sensitivity and specificity with the second reviewer more accurately identifying patients with pain than the first, with sensitivity of 0.922 (95% CI: 0.915, 0.929) and 0.959 (95% CI: 0.953, 0.964), respectively (difference= -0.037, p <0.001). Reviewers were found to have similar specificity of 0.948 (95% CI: 0.941, 0.954) and 0.956 (95% CI: 0.950, 0.962), respectively (difference= -0.008, p=0.065).

### Prevalence and description of patients arriving in pain

Based on the documentation of the triage nursing assessment, 5,509 (55.22%, 95% CI 54.24%, 56.20%) of patients presenting to the ED had signs / symptoms of pain on arrival. The population’s demographics presenting with and without signs of pain on arrival are described in Table 2. Patients with pain were younger, more likely to be born overseas, had a higher socioeconomic status, walked in, and were seen in a lower acuity area of the department and discharged home or to the ED short stay unit.

**Table 2:**
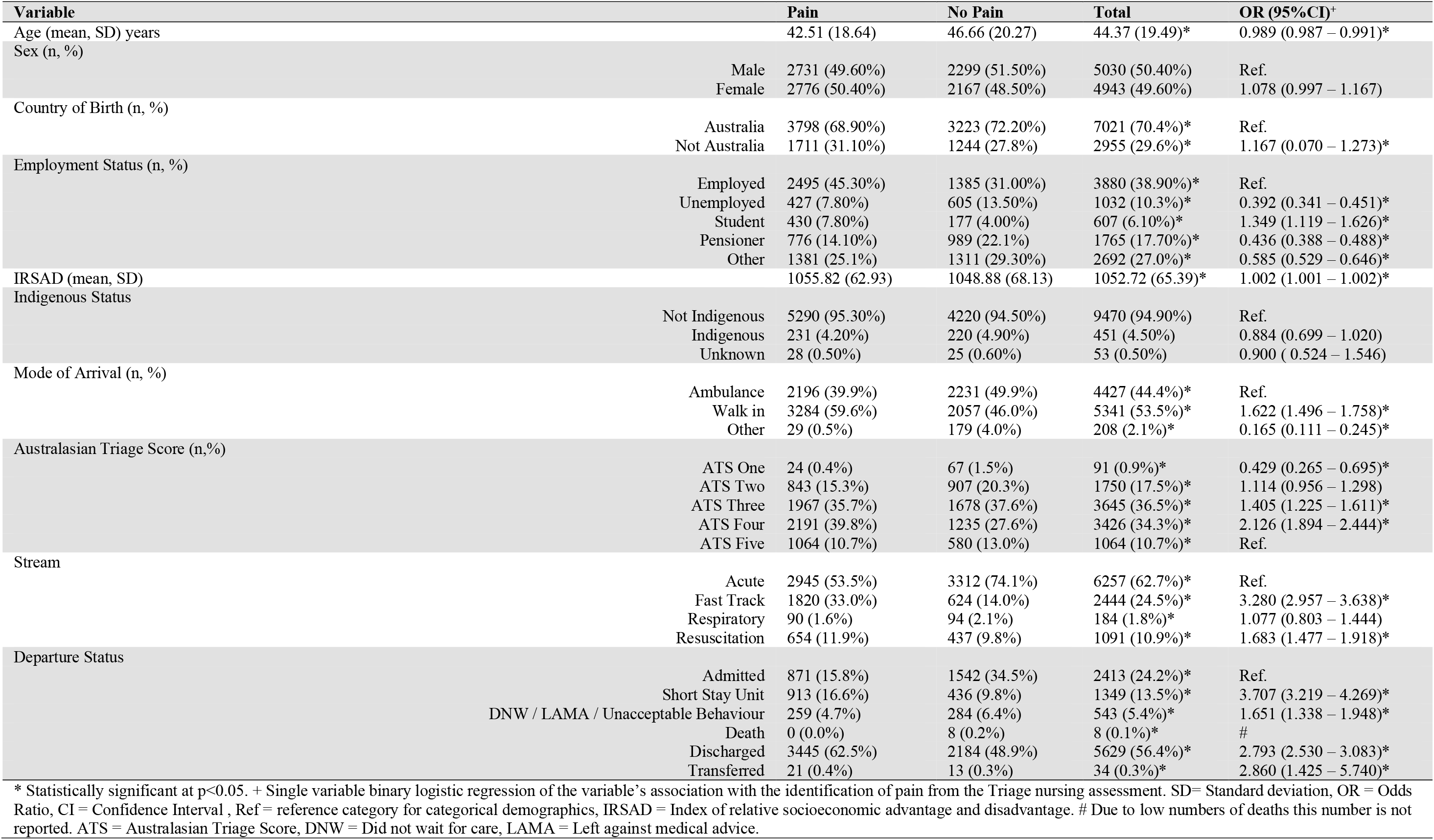
Description of the patient demographics and arrival characteristics of patients presenting with/out signs of pain in their triage nursing assessments

There were minimal variation in painful presentations over the week, with only Sunday having more painful presentations than the rest of the week (OR 1.273, 95% CI 1.098, 1.477). Patients who presented in pain waited longer to be seen by a Provider than a patient that did not present in pain (50.42 (SD 48.83) minutes vs 47.96 (SD 47.41) minutes, p=0.015). However, when considering overall ED length of stay, patients presenting in pain stayed in the ED on average 12.31 (SD 4.08) minutes less (p=0.003). Patients who presented with signs of pain were had reduced odds of representing within 72 hours (OR 0.716, 95% CI 0.589 – 0.871, p<0.001)

### Pharmacological Treatment Provided

A total of 2,511 (45.6%, 95%CI 44.26%, 46.90%) of all patients identified in pain had an analgesic administered in the ED in a median time to the first analgesic of 65 minutes (IQR 38 – 114 minutes). Patients who received analgesia received an average of 2.46 doses (range 1 – 18). Table 3 identifies the most common analgesic dispensed. Table 4 describes the differences between patients identified in pain who received analgesia and those who did not. Table 4 also presents the Hazard ratio for each of the variables associated with the time to first analgesic medication taking into account the observed time (EDLOS) for patients who did not receive analgesia.

**Table 3:** Top 10 dispensed analgesic medications

| **Medication** | **Frequency** | **Percent** |
| --- | --- | --- |
| Acetaminophen (Paracetamol) | 1704 | 27.56% |
| Oxycodone | 1692 | 27.37% |
| Ibuprofen | 924 | 14.95% |
| Fentanyl | 624 | 10.09% |
| Morphine | 342 | 5.53% |
| Pantoprazole | 128 | 2.07% |
| Aspirin | 101 | 1.63% |
| Ketorolac | 61 | 0.99% |
| Hyoscine-N-Butylbromide | 52 | 0.84% |
| Oxycodone - Naloxone 10mg-5mg | 42 | 0.68% |

**Table 4:**
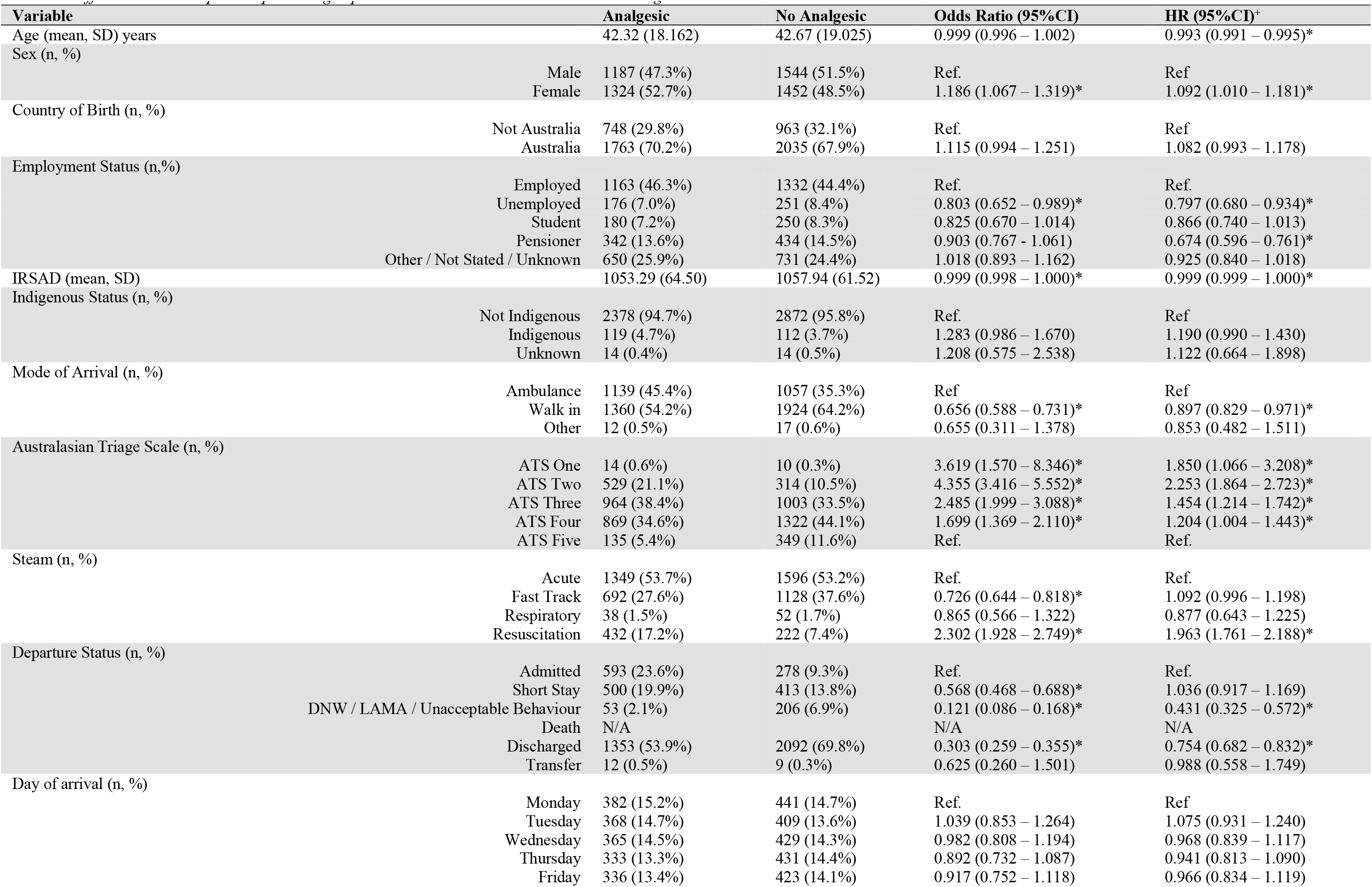
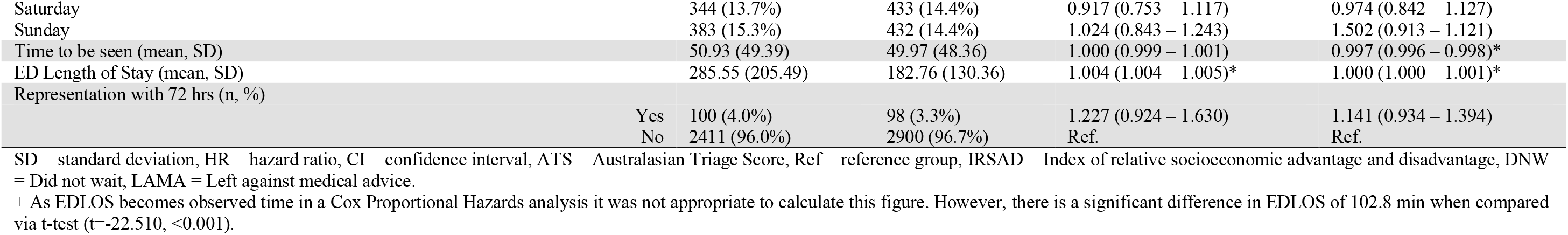
Differences between patients presenting in pain who received and did not receive an analgesic medication.

However, 713 (16.0%) of patients who did not have pain identified at triage had an analgesic medication administered in a median time of 108 minutes (IQR 59 – 219.5 minutes). These patients waited significantly longer for analgesic medications (HR 0.270, 95% CI 0.248, 0.294, p<0.001). They may represent a cohort of patients who did have pain during their ED stay and were not identified at triage. If we were to consider these patients to have signs and symptoms of pain (by the administration of analgesic medication), then the overall proportion of patients experiencing pain during their ED stay increases to 62.37% (95% CI 61.41%, 63.32%).

## Discussion

We have presented a retrospective chart review method to identify patients presenting in pain to the ED using a binary classification of the triage nursing assessment. Using this method, we have identified that 55.22% (95% CI 54.24%, 56.20%) of all patients presenting to the ED of the RBWH between March 2018 and February 2021 had pain on arrival. The processes used in this work have been adapted from previous methods (1, 9); however, its application to triage triage nursing assessments has only been described twice previously in the same patient sample by the lead author (18, 40). The rigour introduced into this method has increased the between reviewer agreement from a Kappa of 0.745 (95%CI 0.716, 0.774) (18) to 0.807 (95%CI 0.787, 0.826). The reviewers had a similar high specificity (0.948 vs 0.956, p=0.065), and a slightly different sensitivity (0.922 vs 0.959, p<0.001). This represents a natural variation in clinical opinion. Nevertheless, the method described is reproducible in many settings, with the documentation of triage nursing assessment being standard emergency care worldwide (47). In addition, the instructions that were given to the abstractors in this work (contained within supplementary material) are adaptable to local workflows and languages, increasing the method’s reproducibility.

The prevalence identified using this method in this population (55.22%, 95%CI 54.24%, 56.20%) compares well with previously reported prevalence using similar methods (56.0% - 61.2%) (1, 8, 9) and with pain prevalence identified in a similar population (59.0%) (20). Variations seen in the prevalence reported may be linked to the methods used, including the inclusiveness of this sample, the lack of reliance on self-report (20) and variations in case-mix seen between studies conducted in Hong Kong (9) and the United States (1). In New Zealand figures that most closely replicate the Australian population (8), there is a significant overlap of the confidence intervals – indicating no statistical differences between the New Zealand prevalence and reported here.

The cohort identified with pain on arrival differed from those without pain on arrival in terms of age, country of birth, measures of socioeconomic status (IRSAD and employment status), mode of arrival, acuity (ATS and stream) and departure status. This may represent differences in the prevalence of pain on arrival in these cohorts or may represent an extension of biases shown in pain care within some of these groups. The cohort of patients identified with pain was younger than the group without signs of pain on arrival (mean difference 4.15 years, OR 0.989, 95%CI 0.987, 0.991). Age is a well-known risk factor for poor pain care in the ED, with patients at extremes of age (less than ten years and greater than 65 years) are at risk of poor care in terms of the provision of analgesia (16) and the patient experience of pain care in the ED (48). Therefore, the differences seen in age may result from previously demonstrated bias rather than an actual association between the prevalence of pain on arrival to the ED and age. The patient’s socioeconomic status is a complex, healthcare setting dependant variable that is likely to reflect societal norms. In a universal healthcare system such as Australia, the patient’s socioeconomic status should cease to matter when receiving care in a publicly funded ED. However, previous studies within Australia have shown that differences in the metrics used within this study (IRSAD and Employment Status) have varying effects on pain care. When considering time to analgesia, both IRSAD and Employment status positively affected the time to the first analgesia, with employed patients and those residing in higher SES areas receiving faster analgesic medication (16). However, these variables did not affect the documentation of pain intensity or the experience of pain care in the ED (18, 48). Mode of arrival, acuity and pain are closely linked. Pain is one of the factors considered when determining the triage category, and minor medical emergencies may receive a higher triage category if they are associated with increasing amounts of pain (49), which changes where the patient receives care within the ED. In Queensland, the Queensland Ambulance Service (QAS) is one of the most sophisticated pre-hospital providers globally and capable of extensive pain care before arrival at the ED (50). Therefore, patients arriving by QAS are likely to have had their pain treated prior to arrival. The lower prevalence of pain in this cohort reflects the care provided in the pre-hospital setting.

Pharmacological analgesia remains the mainstay of pain care in the ED (17). Within the cohort presenting in pain, 45.6% (95%CI 44.26%, 46.90%) received an analgesic medication in a median time of 65 (IQR 38 – 114) min. The median time to first analgesic medication is similar to other figures reported in similar cohorts in Australia (16, 18, 40, 48, 51). In addition, the range of medications given approximates previously described medications in a similar population with a slight decrease in the overall administration of opioids (47.9% vs 43.7%) however Oxycodone remains the second most used drug for pain in this ED. Female patients who were employed, arrived by ambulance service, and had a higher acuity (stream and triage score) were more likely to receive analgesia. However, when we consider the time taken to deliver this pharmacological analgesia represented by the Hazard ratios in Table 4, age also becomes a factor— older patients wait longer for their analgesia. The influence of patient demographic factors (age and socioeconomic status) in conjunction with components of symptom management strategies (arrival method, triage score and stream) has been previously demonstrated within SMT application to pain care in the ED (16). Health service outcomes such as time to be seen and EDLOS were influenced by the presentation in pain and the speed in which analgesia was provided. These relationships were small and have been previously described (16, 40).

There were 713 (16.0%) patients who received analgesic medication in the ED, however, they were not identified as presenting in pain. In addition, the time to first analgesic medication was significantly longer in this group (median of 108 vs 65 minutes), indicating that this cohort of patients either, had pain that was not recognised or documented, developed pain in the ED, care provided caused pain or adequate pre-hospital treatment wore off during their stay. Previous authors have included these numbers in the prevalence reported (5, 6, 9). However, if this figure is included, the prevalence changes from pain on presentation to pain at any stage during the ED stay.

## Limitations

There are several limitations of the method described in this work. First, the retrospective chart review methodology relies on the clinician to document or indicate that the patient has presented with signs or symptoms of pain. We know from previous work that compliance is low when we rely on clinicians to document pain intensity. While this study presents methods that are more robust than simple pain intensity scoring because it is not reliant on standardised documentation, it is still open to the bias of the clinician. The use of clinician abstractors also presents the opportunity for bias. Clinician abstractors interpreting other clinician documentation may reinforce unconscious bias prevalent in this community of practice. However, using three abstractors interpreting hundreds of clinician documentation should, in theory, reduce any individual unconscious bias but reinforce widely shared bias within this setting. Using written instructions has increased the inter-rater reliability of the abstraction. However, the use of abstraction methods that can either reduce bias or identify bias may further improve the identification of patients in pain.

There are limitations of the data used for this study. First, while the missing data was limited and predictable, it was still present within the dataset. There is the possibility that medications were given to patients that were not dispensed via the Pyxis™ system. While medications such as opioids are tightly controlled and always dispensed from Pyxis™, other medications such as Acetominophen and Ibruprofen may be dispensed from other sources, influencing the time to first analgesia. Finally, this study looks at one defined part of the continuum of care. For many patients, pain care will commence before arrival to the ED and continue post-discharge or admission.

## Future Work

Two things need to happen to progress this work and further refine the method described. First, the reproducibility of this method into other EDs within Australia and in other countries needs to be tested, and there needs to be a further reduction in the bias associated with having human abstractors. Currently, this specific method has only been used in one department and, while theoretically, this method should work for any ED, this has yet to be tested. However, replication studies and the use of this method to identify cohorts for the evaluation of interventions will inform the generalisability of the method and set a precedent for acceptable inter-rater reliability. The second area for future work is to reduce the bias inherent in the method through human abstractors. Previous studies of symptoms in specific populations (32, 33) have used NLP, a method stemming from AI to identify free text in medical records that indicate the presence of symptoms. This methodology has been piloted by the authors of this study (34, 35). They have achieved 91% accuracy through deep learning models compared to human abstractors (34). With further refinements in the method described within this work and a more extensive training dataset, it is hoped that this can be used to further improve “pain” classification models and one that is robust to unseen triage nurse assessments. The second benefit of using the AI approach is that it removes the limitations on sample size. The processing of population-level data becomes a viable option. Algorithms may also be deployed in real-time to assist clinicians in making decisions and evaluating interventions(52). However, with the current work, we have developed a basis for vocabulary and gold standard classification to apply NLP and build both machine and deep learning models to complete the classification task at scale.

## Conclusions

This work presents a detailed method for identifying patients who present to the adult ED in pain. This methodology is built upon the previously described retrospective chart review methodology. However, it is unique in its use of triage nursing assessment. Using this methodology, we have identified a prevalence of pain on arrival to a large inner-city ED of 55.2% in a representative sample of 10,000 patients presenting over three years. This prevalence compares well to other settings and similar methodologies. The identified population presenting in pain differed from those not in pain in terms of age, country of birth, socioeconomic status, mode of arrival, urgency and departure destination. There are limitations to the methods that are presented. However, some of these methods can be overcome by using AI, especially NLP, in future iterations. Future work should concentrate on building AI-based approaches to identifying pain in the adult ED.

## Supporting information

Supplimentary Material One

Supplimentary Material Two

Supplimentary Material Three

## Data Availability

All data produced in the present study are available upon reasonable request to the authors

### Appendix

**Appendix A:** Comparison between the sample and population.

| Variable |  | Population | Sample | p-value |
| --- | --- | --- | --- | --- |
| Age (mean, SD) years |  | 44.14 (19.42) | 44.37 (19.49) | 0.245 |
| Sex (n, %) | Male | 112745 (49.94%) | 5030 (50.44%) | 0.328 |
|  | Female | 113033 (50.06%) | 4943 (59.56%) |  |
| IRSAD (mean, SD) |  | 1052.79 (64.53) | 1052.72 (65.39) | 0.926 |
| Mode of Arrival (n, %) | Ambulance | 100266 (44.41%) | 4427 (44.38%) | 0.487 |
|  | Walk in | 120434 (53.33%) | 5341 (53.54%) |  |
|  | Other | 5113 (2.26%) | 208 (2.09%) |  |
| Australasian Triage Score (n, %) | ATS One | 2180 (0.97%) | 91 (0.91%) | 0.711 |
|  | ATS Two | 38712 (17.14%) | 1750 (17.54%) |  |
|  | ATS Three | 82116 (36.36%) | 3645 (35.54%) |  |
|  | ATS Four | 78824 (34.91%) | 3426 (34.34%) |  |
|  | ATS Five | 23981 (10.62%) | 1064 (10.67%) |  |
| Daily time to be seen (mean, SD) |  | 48.17 (13.81) | 47.98 (13.78) | 0.181 |
| Daily ED length of stay (mean, SD) |  | 231.56 (32.55) | 231.06 (32.81) | 0.134 |
| Daily census (mean, SD) |  | 231.29 (23.15) | 231.29 (23.18) | 0.982 |

SD – Standard Deviation, IRSAD – Index of relative socioeconomic advantage or disadvantage, ATS – Australasian triage score, ED – Emergency Department.

