## Supplementary material for "Identifying patients presenting in pain to the adult emergency department: A binary classification task and description of prevalence": Supplimentary Material One

*Supplementary Table One: Definitions of the variables collected and reported within this study*

| Variable | Symptom Management Theory Domain | Definition |
|---|---|---|
| Age | Person | Age of the patient on presentation to the ED. Calculated as the patients date of arrival in the ED minus their date of birth. |
| Sex | Person | Self-identified sex of the patient on arrival. |
| Country of Birth | Person | Self-identified country of birth reduced to a binary categorical variable for the purposes of this study as either Australia or not Australia. |
| Indigenous Status | Person | Does the patient self-identify as an Aboriginal or Torres Straight Islander Australian |
| Australasian Triage Score (ATS) | Components of symptom management strategies | A measure of urgency to be seen by a medical officer on presentation to the ED (Forero & Nugus, 2012). Score is determined by a trained triage Nurse after an initial assessment. |
| Employment Status | Person | The self-reported employment status of the patient on presentation to the ED. |
| Index of relative socio-economic advantage and disadvantage (IRSAD) | Person | A quantitative measure of the relative advantage or disadvantage of the postcode in which the patient lives based upon answers to the previous census (Australian Bureau of Statistics, 2016) |
| Stream | Environment | The location of the ED in which the patient first received care. This indicates the level of urgency, complexity and resource requirements based upon the patient presentation and is decided by the Triage nurse. |
| Mode of Arrival | Components of symptom management strategies | How the patient arrived at the ED. This can either be by ambulance service, by private transport or other methods which include Police service, community transport, mental health care coordinator or prison transport. |
| Departure Status | Component of symptom management strategies | The location in which the patient was discharged. Typically home, to the short stay unit, or admitted to the hospital or transferred to another facility. |
| Analgesia Received | Outcome | Did the patient receive a medication aimed at relieving the patient’s pain while in the ED (defined in supplementary material two). |

| | | |
|---|---|---|
| Time to First Analgesic Medication | Outcome | The time is taken to deliver analgesia, from the triage time to the time of administration of the first medication aimed at relieving the patient's pain. For the purposes of this study the time that the medication was dispensed from Pyxis™ is considered the time of administration. |
| Emergency Department Length of Stay (EDLOS) | Environment | The total length of time that the patient spends in the ED, from time of triage to actual departure. This does not include time spent in the ED short stay unit. Measured in minutes. |
| Time to be seen by a Provider | Environment | The time taken for the patient to be assessed by a Medical Officer or Nurse Practitioner. Measured in minutes. |
| Census | Environment | The total number of patients presenting for care to the ED over a 24 hour period. |
| Day of arrival | Environment | The day of the week in which the patient presented to the ED for care. |
| Representation | Outcome | Did the patient represent within 72 hours of discharge from the ED. Presented as a binary classification. |

ED = Emergency Department.

Australian Bureau of Statistics. (2016). *Socio-Economic indexes for areas* Retrieved from https://www.abs.gov.au/ausstats/abs@.nsf/Lookup/by%20Subject/2033.0.55.001~2016~Main%20Features~IRSAD~20

Forero, R., & Nugus, P. (2012). Australasian College for Emergency Medicine (ACEM) literature review on the Australasian triage scale (ATS). *Institute of Health Innovation*.
