## Supplementary material for "Identifying patients presenting in pain to the adult emergency department: A binary classification task and description of prevalence": Supplimentary Material Two

Supplimentary Material Two: Definition of Analgesic Medication

The definition of an Analgesic Medication is complex when the broad range of medications that may be prescribed to relieve pain are considered. In Emergency Care, a small group of analgesic medications make up the majority of all medications used to relieve pain. For example, in recent work in the Emergency Department, Paracetamol, Oxycodone, Ibruprofen, Morphine, and Fentanyl made up 93.8% of all first-line analgesic medications used (1). However, many medications with analgesia not considered their primary indication may induce analgesia in specific conditions (2). An example of this would be Chlorpromazine, a typical antipsychotic, in headaches and migraines (3). Therefore, several classification methods have been proposed for analgesic medication, including clinical efficacy, therapeutic class, mechanistic approaches, or simply those medications listed on the World Health Organisation's Pain Ladder (2).

In the period that this study was conducted, there were 354 unique medications dispensed from the electronic dispensing system located within the emergency department. Identifying which medications were used for their analgesic properties is essential in this study as outcomes measures such as time to first analgesic medication, the receipt of analgesic medication, and the receipt of an opiate medication are being calculated. Therefore, we propose that the medication dispensed in the emergency department be categorised as follows for the purposes of this study:

1. *Analgesic Medication:* A primary medication to relieve pain and is expected to relieve pain in a wide variety of conditions presenting to the emergency department. All, or almost all use of this medication in the Emergency Department is expected to be used for analgesia.
2. *Local Anaesthetic:* A medication that modulates the pain sensation and may be used a) topically or b) parentally either by local infiltration or systemic administration.
3. *Drugs used to treat pain for specific conditions:* Medications generally not considered analgesic medications; however, they may be an analgesic medication when used to treat a specific condition. These may be considered a) first-line agents or b) second-line medications.
4. *Non-analgesic medications*: Medications that are not used to induce analgesia

The clinician members of the research team reviewed all medications. Initially, JH, AS, and KC sorted medications into Analgesic and Non-analgesic Medications. On further advice from KC and RJ, medications were then separated into the four categories presented. Finally, all clinician members of the research team (JH, KC, RJ, CD and AS) reviewed the final categorisation and reached a consensus.

Supplimentary Material Two: Definition of Analgesic Medication

Time to first analgesic medication will be defined as the time of arrival (triage time) until the first (1) Analgesic Medication or (2) Local Anaesthetic or (3) First Line Drugs used to treat pain for specific conditions is dispensed from the electronic medication system.

The following table (Supplimentary Table 2) identifies the medications categorised as an *Analgesic Medication* or a *Local Anaesthetic*. Table 2 identifies the medications that were categorised as *Drugs used to treat pain for specific conditions* and the specific conditions used to treat. Finally, table 3 identifies the medications that were identified as *Non-analgesic medications.*

*Supplementary Table 2:* List of medication categorised as Analgesic Medication and Local Anaesthetic Medication

| **Analgesic Medication** | **Local Anaesthetic:** |
|---|---|
| Aspirin | Bupivacaine |
| Aspirin - Codeine | Bupivacaine - Adrenaline (Epinephrine) |
| Buprenorphine | Cinchocaine - Zinc Oxide |
| Codeine Phosphate | Lidocaine (Lignocaine) |
| Fentanyl | Lidocaine (Lignocaine) - Phenylephrine |
| Ibuprofen | Lidocaine (Lignocaine) - Prilocaine |
| Indomethacin/Indometacin | Lidocaine (Lignocaine) – Adrenaline (Epinephrine) |
| Ketamine | Ropivacaine |
| Ketorolac | |
| Mefenamic acid | |
| Morphine | |
| Oxycodone | |
| Oxycodone - Naloxone | |
| Paracetamol (Acetominophen) | |
| Paracetamol (Acetominophen) - Codeine | |
| Tapentadol | |
| Tramadol | |

Supplimentary Material Two: Definition of Analgesic Medication

*Supplimentary Table 3:*Drugs used to treat pain for specific conditions

| Medication | Indication | First Line / Second Line |
|---|---|---|
| Aluminium Hydroxide - Magnesium Hydroxide | Epigastric Pain | First Line |
| Baclofen | Muscle Spasm | Second Line |
| Chlorpromazine | Headache | Second Line |
| Clonidine | Various Painful Conditions | Second Line |
| Colchicine | Gout | Second Line |
| Diazepam | Musculoskeletal pain | Second Line |
| Droperidol | Headache | Second Line |
| Famotidine | Epigastric Pain | Second Line |
| Glyceryl Trinitrate (GTN) | Chest Pain | First Line |
| Hyoscine-N-Butylbromide | Abdominal Pain | First Line |
| Isosorbide Mononitrate | Chest Pain | First Line |
| Metoclopramide | Headache | First Line |
| Ondansetron | Headache | First Line |
| Pantoprazole | Epigastric Pain | Second Line |
| Prazosin | Renal Colic | Second Line |
| Pregabalin | Chronic and Neuropathic Pain | Second Line |
| Prochlorperazine | Headache | Second Line |
| Promethazine | Headache | Second Line |
| Propofol | Headache | Second Line |
| Sodium Citrotartrate | Urinary Pain | Second Line |


1. Hughes JA, Alexander KE, Spencer L, Yates P. Factors associated with time to first analgesic medication in the emergency department. Journal of Clinical Nursing. 2021.
2. Lussier D, Beaulieu P. Overview of Pain Management. Adjuvant Analgesics. 2015:1.
3. Shao E, Hughes J, Eley R. The presenting and prescribing patterns of migraine in an Australian emergency department: A descriptive exploratory study. World journal of emergency medicine. 2017;8(3):170.
