## Supplementary material for "Identifying patients presenting in pain to the adult emergency department: A binary classification task and description of prevalence": Supplimentary Material Three

Thank you for agreeing to participate as a Clinical Expert in this study. The aim of this study is to create an artificial intelligence tool for retrospectively identifying patients with pain on presentation to the emergency department. To do this we need a training dataset of presentations that have been manually coded by clinical experts. In this task you (the clinical expert) will be provided with data in an Excel spreadsheet from a number of presentations, and you are required to classify each presentation as either 'Pain' or 'No pain' based on the information available in the triage assessment and whether it indicates that the patient had pain on arrival.

In the triage assessment, pain may be indicated by either a documented pain intensity score such as "pain 9/10 on arrival", or by a verbal descriptor such as "patient states mild pain in abdomen". In some cases, the clinical expert may use their clinical knowledge and experience to determine whether or not the triage assessment indicates that the patient has presented in pain. Presentations in which the triage assessments do not indicate pain or specifically document that the patient has no pain are classified as "No pain". Specific terms that indicate pain include: burning, aching, tearing, throbbing, headache, deformity, or variations thereof.

Here are some examples of triage assessments and the associated "Pain" or "No Pain" classification (highlighted words and phrases indicate pain).

| Presenting Problem | Presenting Problem Nursing Assessment | Pain / No Pain |
|---|---|---|
| 8/10 HYPOGASTRIC PAIN WORSENING SINCE 2100 // WORSE ON MOVEMENT AND PALPATION // NO N&V // NO URINARY SYMPTOMS | ABCD GROSSLY INTACT | Pain |
| GCS 10. INTENTIONAL POLYPHARM OD// ?? OLANZ + DIAZ+ FLUOXETINE + PARACETAMOL | HR 100// HYPOTENSIVE 90 SYS. | No Pain |
| 4-5/7 ABDO CRAMPING + FREQUENT DIARRHOEA + DECREASED PO INTAKE // FEBRILE 38.8 | "OTHER OBS WNL.<br>NIL SIG MED HX<br>NIL REG MEDS" | Pain |
| INVERSION INJURY R) ANKLE DURING FOOTBALL, UNABLE TO WEIGHT BEAR, SWELLING LATERAL SIDE | UNABLE TO WEIGHT BEAR, NIL PARESTHESIA, GOOD WARMTH AND COLOUR | Pain |
| 5/7 SOBOE PROGRESSIVLEY WORSENING, INCREASING FATIGUE, PRODUCTIVE COUGH WITH CHANGE IN COLOUR OF SPUTUM HX: COPD, T2DM | "SPO2 94% ROOM AIR WITH QAS, BSL 13.9MMOL, QAS REPORT COARSE CRACKLES AND EXP WHEEZE REFUSED VENTOLIN WITH QAS, HAS TAKEN OWN ATROVENT WITH LITTLE IMPROVEMENT REPORTED BY PT, QAS: NIL MEDS, HR~86 ON MONITOR, QAS REPORT 12 LEAD ECG NAD BUT NOT VISUALISED B" | No Pain |
| HEADACHE, VOMITING, FEVER | DENEIS OS TRAVEL, NO COUGH, NO OTHER MEDICAL COMORBIDS | Pain |
| ETOH (2 BOTTLES VODKA). EMOTIONAL. URINARY/FAECAL INCONTINENT. | A - PATENT B - NIL RESP DISTRESS C - RAD PULSE STRONG D - GCS15. OBS WNL W. QAS. NOT FORTHCOMING WITH INFORMATION. | No Pain |
| LARGE LACERATION TO L ANKLE/HEEL FROM KITCHEN KNIFE | "A=PATENTB=SOT C=STRONG REG PULSE, HR 88 D= GCS 15PT HAS COVERED WITH DRESSING. ATTEMPTED TO UNCOVER TO VIEW WOUND AT TRIAGE BUT TO PAIFUL FOR PT. HAEMOSTASIS. ABLE TO SEE LARGE LACERATION BUT UNABLE TO ASSESS SIZE OR DEPTH DUE TO LARGE CLOT. PT REPORTS" | Pain |

Access your Excel spreadsheet via the link provided. For each presentation (row), review the triage assessment, then select "Pain" or "No pain" from the drop-down list. We recommend that you perform the task for no longer then 45 min at a time, to avoid fatigue that can impair your ability to accurately classify the presentations. Take regular breaks and do only small amounts of classification at any time. Once you have finished the classification task, please let the Principal Investigator know:
